# Forecasting U.S. 12-Month-Ending Overdose Death Counts: Multi-Model National and Regional Projections

**DOI:** 10.64898/2026.08.03.26359533

**Authors:** Faharudeen Alhassan, Hamed Karami, Robert Bohler, Isaac C. H. Fung, Svenn-Erik Mamelund, Sunmi Lee, Emily N. Peterson, Gerardo Chowell

## Abstract

**Aims:** To assess whether recent declines in U.S. rolling 12-month-ending drug overdose death counts are projected to continue, compare the retrospective performance of short-term forecasting models, and estimate national and regional 12-month changes.

**Design:** Comparative time-series forecasting study with a retrospective March 2025–February 2026 forecast evaluation and subsequent 12-month-ahead projections through February 2027.

**Setting:** United States and four U.S. Census regions: Northeast, Midwest, South, and West.

**Cases:** Aggregate drug overdose deaths reported in the National Center for Health Statistics Vital Statistics Rapid Release system (VSRR) and identified using ICD-10 underlying cause-of-death codes X40–X44, X60–X64, X85, and Y10–Y14.

**Measurements:** The primary outcome was the monthly series of rolling 12-month-ending overdose deaths. Candidate models included ARIMA, generalized additive models, Prophet, and AICc-ranked *n*-sub-epidemic models. Models were calibrated using January 2020–February 2025 data and evaluated against March 2025–February 2026 observations using mean absolute error, mean squared error, empirical 95% prediction interval coverage, and weighted interval score (WIS). Individual models were ranked by retrospective WIS, and normalized inverse-WIS weights were used to construct ensembles from the top-ranked models. Final forecasts were generated for March 2026– February 2027 after recalibrating models using January 2020–February 2026 data.

**Results:** Retrospective WIS performance differed geographically: GAM performed best nationally and in the Midwest and West, the leading n-sub-epidemic model in the Northeast, and ARIMA in the South. Median forecasts from all individual models and ensembles projected declines from February 2026 to February 2027 nationally and in each region, although prediction intervals varied substantially. Individual national median projections ranged from declines of 12.8% to 25.8%, while weighted-ensemble median projections indicated declines of 20.5% to 21.7%. Projected weighted-ensemble declines were larger in the Northeast (25.4%–30.0%) and Midwest (24.3%–26.3%) than in the South (16.6%–19.5%) and West (18.0%–21.8%).

**Summary:** Ensemble forecasts of the reported provisional VSRR series were consistent with continued declines in rolling annual overdose death counts nationally and across U.S. Census regions. The projected magnitude of decline differed by region and model specification. Because the forecasts used provisional rolling 12-month-ending counts, they should be interpreted as surveillance projections rather than exact monthly mortality predictions.

## 1 INTRODUCTION

After more than two decades of increases in U.S. drug overdose mortality [1], deaths accelerated during the COVID-19 pandemic, coinciding with disruptions in health care access, harm-reduction services, social support, and illicit drug markets [2]. Recent surveillance analyses suggest that U.S. overdose mortality has begun to decline [3, 4]. Mortality nevertheless remains historically high, and it is uncertain whether the decline will persist or occur consistently across geographic areas. Quantifying the near-term trajectory is important for interpreting the epidemic and informing prevention, treatment, and harm-reduction planning.

Jalal et al. described a long-run exponential trajectory in U.S. overdose mortality from 1979 through 2016; Jalal and Burke later discussed shorter-term accelerations and decelerations around that broader pattern [5, 6]. U.S. overdose mortality fell below the Jalal–Burke exponential trajectory in 2024 [7], but whether this marks a sustained reversal or a temporary deviation remains unresolved.

Short-term probabilistic forecasts can help quantify the trajectories implied by recent surveillance data while characterizing uncertainty and assessing whether conclusions depend on model structure. A growing literature has applied statistical, Bayesian, mechanistic, and machine-learning approaches to overdose prediction across national, state, county, and local scales [8–12]. Related work has also used system dynamics, compartmental, and Markov models, including the SOURCE model, to project opioid-related harms and evaluate potential intervention strategies [13–18]. However, it remains unclear how alternative probabilistic model classes compare when applied to the same recent provisional surveillance series, whether performance differs by geography, and whether performance-weighted ensembles provide a less model-dependent summary.

National overdose mortality trends can obscure substantial heterogeneity arising from differences in drug supply, fentanyl penetration, polysubstance use, access to treatment and harm-reduction services, socioeconomic conditions, and reporting practices. Recent national evidence indicates that the 2023–2024 decline was driven largely by reductions in fentanyl-involved deaths, both with and without stimulant involvement. However, deaths involving stimulants without fentanyl and xylazine-involved deaths accounted for growing proportions of overdose fatalities, while substantial racial/ethnic disparities persisted [4]. Geographic heterogeneity is also evident: overdose deaths rebounded in Arizona in early 2025 despite the national decline in 2024 [19], and state-level analyses documented substantial geographic and racial/ethnic variation in excess overdose mortality from 2020 through 2023 [20]. Together, these findings reinforce the need for geographically stratified surveillance. Although state- and county-level forecasts are valuable for local planning, Census-region forecasts provide an intermediate surveillance scale that can summarize broad geographic patterns while avoiding some of the instability associated with smaller-area time series.

In this study, we evaluated whether recent declines in reported U.S. 12-month-ending overdose death counts are projected to continue through February 2027 nationally and across the four U.S. Census regions: Northeast, Midwest, South, and West [21]. To support this aim, we compared the retrospective probabilistic performance of autoregressive integrated moving average, generalized additive, Prophet, and n-sub-epidemic models under a common evaluation design and constructed region-specific performance-weighted ensembles. We asked whether model classes agreed on the direction and magnitude of near-term change, whether the best-performing model differed by geography, and whether the magnitude of projected decline varied across regions.

## 2 METHODS

### 2.1 Data source, outcome definition, and geographic aggregation

We obtained reported monthly 12-month-ending provisional drug overdose death counts from the National Center for Health Statistics (NCHS) Vital Statistics Rapid Release (VSRR) system, which provides provisional overdose death counts based on the current flow of mortality data in the National Vital Statistics System [22]. Drug overdose deaths were identified using underlying cause-of-death ICD-10 codes X40–X44, X60–X64, X85, and Y10–Y14, consistent with standard NCHS and CDC overdose surveillance definitions [22, 23].

Data were extracted on July 16, 2026. Values through February 2026 reflect the reported provisional 12-month-ending counts available on that extraction date and are subject to revision. The February 2026 value was used as the most recent available 12-month-ending count, and no additional completeness correction was applied. Because VSRR counts are updated as additional death certificates, toxicology, and cause-of-death information become available, forecasts should be interpreted relative to the surveillance data available at the time of model calibration [22].

The primary outcome was the 12-month-ending overdose death count, defined as the cumulative number of overdose deaths occurring during the 12 months ending in a given calendar month. We modeled this series directly because it matches the form in which the VSRR surveillance data are reported and provides a smoother, more stable summary of annual mortality trends across regions. Consequently, forecasts and retrospective performance metrics should be interpreted as projections and evaluations of the rolling annual surveillance series rather than independent monthly mortality outcomes.

We analyzed five geographic series: the national series and four regional series corresponding to the U.S. Census regions (Northeast, Midwest, South, and West) [21]. The national series was obtained directly from the NCHS VSRR data source and was not constructed by summing the regional series. Regional series were constructed by aggregating state-level overdose death counts according to U.S. Census Bureau regional assignments. The District of Columbia was included in the South, and Puerto Rico was excluded. The VSRR data report New York State and New York City as separate jurisdictions, with the New York State record excluding New York City. Because the Census regional classification assigns states and the District of Columbia rather than separate city reporting jurisdictions, New York State was included in the Northeast, but the separate New York City record was not added to any regional series. Consequently, the sum of the four regional series does not exactly equal the directly reported national series. This aggregation was used to examine broad geographic heterogeneity while improving stability relative to smaller-area time series. For numerical stability during model fitting, overdose death counts were divided by 1,000 before estimation and transformed back to the original death-count scale for presentation. This scaling was a numerical transformation only and did not convert counts into rates.

### 2.2 Forecasting design

Candidate models were fitted separately to the national series and to each of the four regional series. The analysis followed a two-stage forecasting design. First, each model was calibrated using data from January 2020 through February 2025 and used to generate 12-month-ahead forecasts for March 2025 through February 2026. These retrospective forecasts were evaluated against the observed data from March 2025 through February 2026. Second, after retrospective evaluation, all available data from January 2020 through February 2026 were used to recalibrate the models, and final 12-month-ahead forecasts were generated for March 2026 through February 2027.

This design allowed us to conduct a 12-month retrospective evaluation before producing the final forecasts. Because the outcome was a rolling 12-month-ending count, the evaluation targets overlapped with months included in the calibration period. Prediction interval coverage was calculated as the proportion of the 12 retrospective targets falling within the nominal 95% prediction interval and was interpreted descriptively because coverage estimates based on only 12 targets are unstable.

### 2.3 Candidate forecasting models

The model set included conventional statistical forecasting approaches and phenomenological growth models selected to capture different features of overdose mortality dynamics, including smooth temporal trends, nonlinear change, plateau-like behavior, and shifts in the pace of mortality increase or decline. The candidate models included autoregressive integrated moving average (ARIMA) models, generalized additive models (GAMs), Prophet models, and top-ranked *n*-sub-epidemic models. Each model was used to generate 12-month-ahead point forecasts and corresponding prediction intervals.

ARIMA models were used to capture temporal dependence, non-stationarity, and serially correlated forecast errors in the overdose mortality series. A non-seasonal ARIMA(*p, d, q*) model can be written as

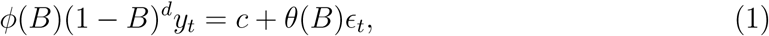

where *y_t_* denotes the scaled overdose mortality series at time *t*, *B* is the backshift operator, *ɛ_t_* is a white-noise error term, *ϕ*(*B*) is the autoregressive polynomial, *θ*(*B*) is the moving-average polynomial, and (1 *− B*)*^d^* is the differencing operator. ARIMA models were fitted using the StatModPredict Shiny application [24] in R version 4.5.1. Seasonal terms were not included. Candidate models were searched over *p* = 0*,…,* 10 and *q* = 0*,…,* 5, with the maximum non-seasonal differencing order set to *d* = 2. The overdose death count series was divided by 1,000 before model fitting for numerical stability, and forecasts were transformed back to the original death-count scale for presentation.

GAMs were used as flexible statistical benchmarks for modeling nonlinear temporal trends. For each geographic series, overdose mortality was modeled as

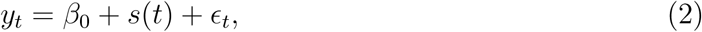

where *y_t_* denotes the scaled 12-month-ending overdose death count at time *t*, *β*_0_ is the intercept, *s*(*t*) is a smooth function of time represented using penalized splines, and *ɛ_t_ ∼ N* (0*, σ*^2^) is the residual error term [25]. GAMs were fitted using the gam function in the mgcv package in R, with a penalized spline smoothing term and normal error distribution. Forecast uncertainty intervals were constructed from the fitted GAM predictions and their standard errors returned by predict.gam with se.fit = TRUE. For each nominal central interval level, lower and upper limits were computed using normal quantiles as *ŷ_t_*+*z_α/_*_2_*SE*(*ŷ_t_*) and *ŷ_t_* + *z*_1_*_−α/_*_2_*SE*(*ŷ_t_*), respectively. The code generated central 10%, 20%,…, 90%, 95%, and 98% intervals; the 95% interval was used for empirical coverage calculations.

Prophet models were used to represent the observed series using a flexible trend-based forecasting framework [26]. In its general form, Prophet decomposes a time series as

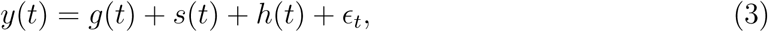

where *g*(*t*) is the trend component, *s*(*t*) represents seasonal patterns, *h*(*t*) represents holiday or event effects, and *ɛ_t_* is the residual error. In this analysis, Prophet was fitted using the prophet package in R with a linear growth trend, with seasonal, holiday, intervention, and external covariate effects disabled. Forecast uncertainty intervals were obtained from Prophet’s model-based uncertainty estimates by fitting the model with multiple interval widths and extracting the corresponding yhat, yhat_lower, and yhat_upper values from the predict function. The code generated central 10%, 20%,…, 90%, 95%, and 98% intervals, with the 95% interval used for empirical coverage calculations.

All models were fitted independently for each geographic series. We used univariate forecasting models so that the comparison focused on the predictive information contained in the overdose mortality surveillance series itself. External predictors, including drug supply indicators, toxicology patterns, treatment access, naloxone distribution, socioeconomic covariates, policy changes, and harm-reduction interventions, were not included. Therefore, the forecasts should be interpreted as projections of recent surveillance trends rather than explanations of the causes of projected regional differences. Future work should evaluate whether incorporating external covariates improves forecast accuracy and interpretation, especially for regions where individual model projections were less consistent.

### 2.4 The *n*-sub-epidemic modeling framework

The *n*-sub-epidemic modeling framework represents complex population-level trajectories as the aggregation of multiple overlapping and asynchronous sub-epidemic components [27–29]. Although originally developed for infectious disease applications, the framework can be used more broadly to approximate nonlinear growth, saturation, plateau-like behavior, and changes in trend in surveillance time series.

In this study, the observed 12-month-ending overdose death count trajectory was modeled as the sum of up to three sub-epidemic components. Let *C*(*t*) denote the cumulative fitted trajectory at time *t*. The aggregate trajectory was represented as

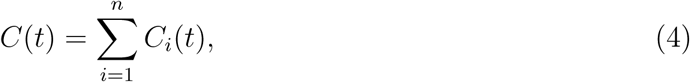

where *C_i_*(*t*) denotes the trajectory of the *i*th sub-epidemic component and *n ≤* 3. Each component followed a generalized logistic growth formulation,

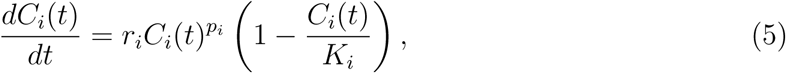

where *r_i_* is the growth-rate parameter, *p_i_* controls the scaling of growth, and *K_i_* is the final size or saturation parameter for the *i*th component. The model allows the observed trajectory to be represented as a combination of nonlinear growth components with different timing, growth rates, and saturation levels.

Candidate *n*-sub-epidemic models were fitted separately for each geographic series. The maximum number of sub-epidemic components was set to three, and onset timing was fixed. Parameter estimation was performed by nonlinear least squares under a normal error structure applied to the model increments. To reduce sensitivity to local minima, each model fit used 20 initial parameter guesses in a MultiStart optimization procedure.

Uncertainty was characterized using bootstrap resampling with *B* = 300 bootstrap realizations. For each bootstrap realization, the model was refitted and used to generate a forecast trajectory over the 12-month forecast horizon. Prediction intervals were obtained from the empirical distribution of the bootstrap forecast trajectories at each forecast time point. The median forecast was defined as the pointwise median of the bootstrap trajectories, and the 95% prediction interval was defined using the corresponding pointwise 2.5th and 97.5th percentiles.

Candidate sub-epidemic structures were ranked using the corrected Akaike information criterion (AICc). The terms Rank 1, Rank 2, and Rank 3 refer only to the first-, second-, and third-best *n*-sub-epidemic candidate models according to AICc within each geographic series. These AICc-based ranks are specific to the *n*-sub-epidemic model class and should not be confused with the WIS-based ranking used later to construct ensemble models across all candidate forecasting models.

### 2.5 Retrospective evaluation and ensemble construction

Retrospective performance over March 2025–February 2026 was assessed using mean absolute error, mean squared error, empirical 95% prediction interval coverage, and weighted interval score (WIS). WIS was the primary criterion for ensemble construction because it evaluates forecast accuracy, interval calibration, and sharpness [30, 31].

For each geographic series, the six individual models were ranked according to their mean retrospective WIS. Lower values indicated better probabilistic forecast performance. This WIS-based ranking was used only for ensemble construction and is distinct from the AICc-based Rank 1–Rank 3 labels assigned within the *n*-sub-epidemic framework.

Weighted ensembles EW*N*, *N* = 2*,…,* 6, combined the top *N* models using normalized inverse-WIS weights,

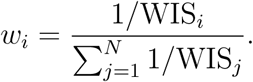

Ensemble medians and prediction limits were calculated as weighted averages of the corresponding model-specific medians and limits. An unweighted ensemble, denoted EU, assigned equal weight to all six models. Weights were estimated separately for each geographic series and applied after the models were refitted using data through February 2026. The weighted average of the component-model WIS values was treated only as a descriptive summary and not as the retrospective WIS of the combined ensemble forecast. Full retrospective model metrics are reported in Table 1, and EW2 inverse-WIS ensemble composition and model weights are reported in Supplementary Table S1.

**Table 1:** Retrospective forecast performance of candidate models by geographic series over the March 2025–February 2026 evaluation period. Within each geographic series, models are ordered by increasing weighted interval score (WIS), so the best-performing probabilistic forecast model appears first. Lower MAE, MSE, and WIS indicate better predictive performance, whereas 95% prediction interval coverage closer to 95% indicates better interval calibration.

| Geographic series | Model | WIS rank | MAE | MSE | 95% PI coverage | WIS |
| --- | --- | --- | --- | --- | --- | --- |
| National | GAM | 1 | 3.53 | 17.29 | 100.00 | 2.00 |
|  | ARIMA | 2 | 4.14 | 23.47 | 100.00 | 2.30 |
|  | Rank 3 | 3 | 11.30 | 127.93 | 16.67 | 7.37 |
|  | Rank 1 | 4 | 10.40 | 108.43 | 0.00 | 7.40 |
|  | Rank 2 | 5 | 12.19 | 148.82 | 0.00 | 7.95 |
|  | Prophet | 6 | 9.86 | 122.92 | 0.00 | 8.25 |
| Northeast | Rank 1 | 1 | 0.50 | 0.38 | 75.00 | 0.34 |
|  | GAM | 2 | 0.83 | 1.00 | 75.00 | 0.52 |
|  | ARIMA | 3 | 1.08 | 1.65 | 100.00 | 0.62 |
|  | Prophet | 4 | 1.60 | 3.41 | 0.00 | 1.35 |
|  | Rank 3 | 5 | 2.52 | 6.38 | 0.00 | 1.69 |
|  | Rank 2 | 6 | 2.93 | 8.63 | 0.00 | 2.17 |
| Midwest | GAM | 1 | 0.17 | 0.04 | 100.00 | 0.29 |
|  | ARIMA | 2 | 0.21 | 0.08 | 100.00 | 0.41 |
|  | Rank 1 | 3 | 0.86 | 1.16 | 66.67 | 0.56 |
|  | Rank 2 | 4 | 2.20 | 4.89 | 91.67 | 1.35 |
|  | Prophet | 5 | 2.03 | 5.27 | 0.00 | 1.71 |
|  | Rank 3 | 6 | 4.02 | 16.35 | 0.00 | 3.18 |
| South | ARIMA | 1 | 2.69 | 9.92 | 100.00 | 1.50 |
|  | Rank 1 | 2 | 2.01 | 6.34 | 50.00 | 1.56 |
|  | GAM | 3 | 2.46 | 8.38 | 50.00 | 1.66 |
|  | Rank 2 | 4 | 2.52 | 9.07 | 50.00 | 1.86 |
|  | Prophet | 5 | 3.52 | 16.42 | 0.00 | 2.89 |
|  | Rank 3 | 6 | 5.03 | 25.34 | 0.00 | 3.39 |
| West | GAM | 1 | 0.20 | 0.09 | 100.00 | 0.21 |
|  | ARIMA | 2 | 0.29 | 0.10 | 100.00 | 0.33 |
|  | Rank 2 | 3 | 2.08 | 4.37 | 83.33 | 1.28 |
|  | Prophet | 4 | 2.28 | 5.92 | 0.00 | 1.87 |
|  | Rank 1 | 5 | 2.48 | 7.33 | 25.00 | 1.89 |
|  | Rank 3 | 6 | 3.11 | 9.84 | 0.00 | 2.17 |

*Note:* WIS rank refers to the retrospective ordering used for ensemble construction. EW2 combines the two best WIS-ranked models within each geographic series, EW3 combines the top three, and so forth up to EW6, which includes all six individual models with inverse-WIS weights. EU is the unweighted ensemble in which all six individual models receive equal weight 1/6. Rank 1, Rank 2, and Rank 3 are individual $n$ -sub-epidemic models selected using AICc within each geographic series and should not be confused with the WIS-based ranks used for ensemble construction.

### 2.6 Projected 12-month percentage change

For each individual model, WIS-based ensemble model, and geographic series, we summarized the final March 2026–February 2027 forecast using the projected percentage change from the last observed value to the 12-month-ahead forecast. This quantity was computed as

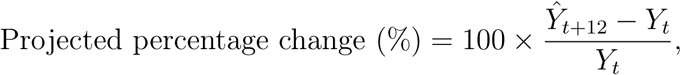

where *Y_t_* denotes the last observed 12-month-ending overdose death count in February 2026 and *Ŷ_t_*_+12_ denotes the forecasted 12-month-ending overdose death count for February 2027. Positive values indicate a projected increase in rolling annual overdose death counts, whereas negative values indicate a projected decline. This summary provides an interpretable measure of the projected near-term direction and magnitude of change and complements the full month-by-month forecast trajectory.

## 3 RESULTS

### 3.1 Retrospective forecasting performance

Retrospective performance differed by model and geographic series. GAM had the lowest WIS nationally and in the Midwest and West, the AICc-ranked *n*-sub-epidemic Rank 1 model performed best in the Northeast, and ARIMA performed best in the South. ARIMA achieved 95% interval coverage for all 12 retrospective targets in each series, whereas Prophet achieved no coverage in any series.

Table 1 summarizes retrospective MAE, MSE, WIS, and empirical 95% prediction interval coverage by geographic series and model. MAE and WIS are reported in thousands of deaths because overdose death counts were divided by 1,000 before model fitting; MSE is reported in squared thousands of deaths. Because coverage was evaluated using only 12 retrospective forecast targets, empirical coverage should be interpreted descriptively.

The WIS-based rankings were used to construct region-specific inverse WIS-weighted ensembles. The weighted average of component-model WIS values was used only to define and summarize the ensemble weights and was not interpreted as the retrospective WIS of the combined ensemble forecast.

### 3.2 Prospective national and regional projections

After recalibrating the models using data from January 2020 through February 2026, we generated 12-month-ahead forecasts through February 2027 and calculated projected percentage changes relative to the reported February 2026 value. All model median forecasts were below the reported February 2026 value, although prediction intervals differed substantially across models.

At the national level, individual model median projections ranged from a 12.8% decline under Rank 3 to a 25.8% decline under Rank 1. ARIMA and GAM projected median declines of 20.2% and 23.0%, respectively, while Prophet projected a 19.0% decline. Weighted ensemble median projections were tightly grouped, ranging from 20.5% under EW3 to 21.7% under EW2, and the unweighted ensemble projected a 20.6% decline.

Regional model median forecasts also indicated declines, but the magnitude varied geographically. In the Northeast, weighted ensemble medians projected declines ranging from 25.4% to 30.0%. In the Midwest, weighted ensemble medians projected declines ranging from 24.3% to 26.3%. In the South, weighted ensemble medians projected declines ranging from 16.6% to 19.5%. In the West, weighted ensemble medians projected declines ranging from 18.0% to 21.8%. Between-model variation was greater in the South and West than in the national, Northeast, and Midwest series.

For a parsimonious illustration of the two-model inverse-WIS ensemble, EW2 projections are summarized in Table 2. Full individual-model and ensemble projections are shown in Figure 1.

**Figure 1:**
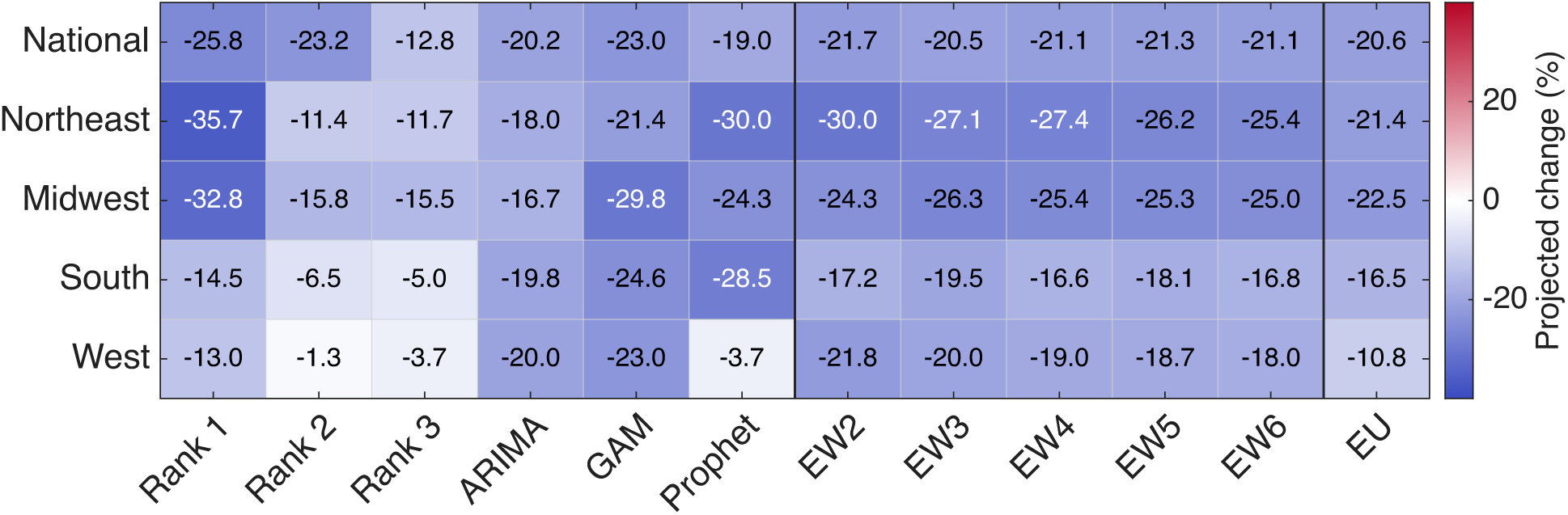
Median projected 12-month percentage change in rolling annual overdose death counts from February 2026 to February 2027 by geographic series and forecasting model. Rows correspond to geographic series and columns correspond to individual and ensemble forecasting models. All displayed median projected percentage changes are negative, indicating projected declines across all geographic series and model classes. Rank 1–Rank 3 denote AICc-ranked individual *n*-sub-epidemic models. EW2–EW6 denote inverse-WIS weighted ensembles constructed from the top 2 through top 6 retrospectively ranked models, and EU denotes the unweighted ensemble using all six individual models.

**Figure 2:**
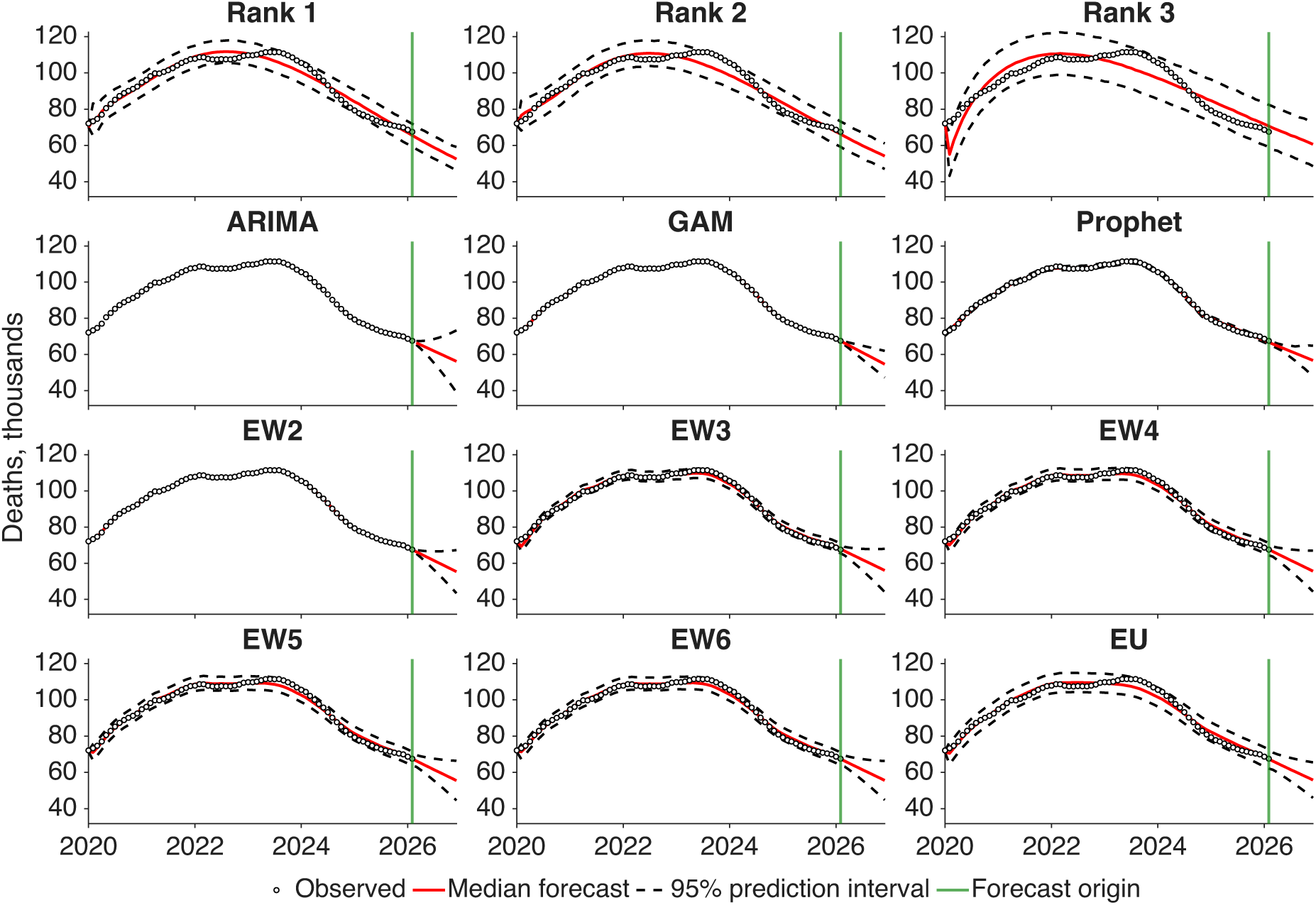
National 12-month-ahead forecasts of rolling annual drug overdose death counts by forecasting model. Observed 12-month-ending overdose death counts from January 2020 through February 2026 are shown as black circles. The vertical green line marks the forecast origin at February 2026, after which models generate forecasts for March 2026 through February 2027. Red curves indicate model-specific median forecasts, and dashed black curves indicate the corresponding 95% prediction intervals. Panels show forecasts from the three AICc-ranked *n*-sub-epidemic models, ARIMA, GAM, Prophet, WIS-based weighted ensembles EW2–EW6, and the unweighted ensemble EU.

**Table 2:** Observed February 2026 12-month-ending overdose death counts and EW2 ensemble-projected February 2027 counts by geographic series.

| Geographic series | Observed Feb 2026 | Model | Projected Feb 2027 | Projected change (%) |
| --- | --- | --- | --- | --- |
| National | 67,531 | EW2 | 52,910 | -21.7 |
| Northeast | 9,121 | EW2 | 6,380 | -30.0 |
| Midwest | 11,707 | EW2 | 8,858 | -24.3 |
| South | 23,689 | EW2 | 19,620 | -17.2 |
| West | 21,130 | EW2 | 16,520 | -21.8 |

## 4 DISCUSSION

In this comparative forecasting study, all individual-model and ensemble median forecasts indicated lower provisional rolling 12-month-ending overdose death counts by February 2027. Weighted-ensemble medians projected a national decline of 20.5%–21.7%, with larger declines in the Northeast (25.4%–30.0%) and Midwest (24.3%–26.3%) than in the South (16.6%–19.5%) and West (18.0%–21.8%). Retrospective performance also varied geographically, and no single model had the lowest WIS across all five geographic series. The consistent direction of the median projections across the model classes examined makes the result less dependent on any one model specification.

In the single-origin retrospective evaluation, GAM had the lowest WIS nationally and in the Midwest and West, ARIMA performed best in the South, and the leading *n*-sub-epidemic model performed best in the Northeast. ARIMA showed the most consistent nominal interval coverage, whereas Prophet’s intervals failed to cover any retrospective targets. These rankings should be interpreted cautiously because they were based on only 12 strongly overlapping targets and may not generalize to other forecast origins.

The magnitude of the median projected decline varied by geographic series. Weighted ensemble medians indicated larger projected declines in the Northeast and Midwest and more moderate projections in the South and West. These regional projections can be interpreted alongside emerging hypotheses about national overdose mortality declines, including changes in fentanyl exposure, drug use, and illicit supply conditions [32, 33]. However, the present models were univariate and cannot attribute regional differences to these or any other mechanisms. The regional results should therefore be interpreted descriptively as model-based projections of recent surveillance trajectories.

The Census regions should not be interpreted as causal or mechanistic units. They are administrative surveillance groupings that combine states with heterogeneous demographic, policy, and drug-market contexts. Their value in this analysis is that they provide an interpretable intermediate scale between national and state-level forecasts. This scale can help summarize broad geographic differences while avoiding some instability associated with smaller-area time series.

The agreement among ensemble median forecasts should also be interpreted cautiously. Individual models differ in their assumptions about temporal dependence, smoothness, nonlinear growth, and trend changes; however, all models were calibrated to the same reported provisional VSRR series and therefore share common information about the recent trajectory. By weighting models according to retrospective WIS, the ensembles reduced reliance on a single model specification while preserving information from multiple plausible trajectories. Nevertheless, model agreement does not provide independent confirmation that the decline will persist, especially because the evaluation used only one forecast origin and 12 overlapping retrospective targets.

These findings are consistent with recent evidence that U.S. overdose mortality has begun to decline after years of substantial increases [3]. National data show that overdose death rates increased markedly over the past two decades but declined from 2022 to 2023, although mortality remained high relative to earlier years [1, 34]. At the same time, the overdose epidemic remains dynamic, shaped by synthetic opioids, stimulant involvement, polysubstance use, and geographic variation in the illicit drug supply [35]. Therefore, projected declines should not be interpreted as evidence that the overdose crisis has resolved. Rather, they indicate that recent reported surveillance patterns, if continued, are compatible with decreasing rolling annual overdose death counts over the next year.

A practical contribution of this study is the use of projected 12-month percentage change as a concise summary of forecasted overdose mortality trajectories. This metric communicates whether rolling annual overdose death counts are projected to increase, decrease, or remain approximately stable over a defined horizon while retaining a clear connection to the underlying forecast trajectory. Beyond this summary measure, the comparative forecasting approach can be updated as new VSRR data become available, uses retrospective WIS-based evaluation to compare model performance, and applies weighted ensembles to reduce reliance on any single model. However, projected percentage change is a point-forecast summary and should be interpreted alongside prediction intervals, retrospective model performance, regional variation, and the known limitations of provisional 12-month-ending mortality counts.

The use of VSRR 12-month-ending counts has important implications for interpretation. Because these counts represent rolling annual totals, the forecasts describe projected changes in smoothed annual overdose death counts rather than incident monthly overdose deaths. This smoothing can improve stability and reduce noise, but it can also attenuate abrupt changes and delay detection of turning points. In addition, the outcome analyzed here was the reported provisional VSRR count available at the time of data extraction, not the eventual final overdose death count. Therefore, differences between projected and subsequently reported values may reflect both true changes in overdose mortality and revisions to provisional surveillance data. Provisional overdose mortality data may be incomplete because overdose deaths often require toxicological testing and death investigation, and the extent of incompleteness can vary by jurisdiction and reporting period [22]. Forecasted declines should therefore be interpreted as projections of the reported provisional surveillance series, conditional on the July 16, 2026 data extraction, and should be updated as revised mortality data become available.

This study has several limitations. First, the analysis used reported provisional VSRR data, and the most recent observations may undergo greater revision than earlier values, potentially affecting the magnitude of the projected declines. Second, the rolling 12-month-ending outcome is highly autocorrelated and may delay detection of abrupt changes. The retrospective targets also overlapped with the calibration period, which may favor smooth or persistent models and influence WIS-based rankings and ensemble weights. Third, empirical coverage of the nominal 95% prediction intervals varied across models; therefore, the intervals should be interpreted as model-based uncertainty bands rather than consistently calibrated predictive intervals. Fourth, the rolling construction reduced the effective amount of independent information, increasing the potential for overfitting and unstable model selection. Fifth, regional aggregation may mask state-level and demographic heterogeneity. The univariate models also excluded external predictors and did not distinguish overdoses involving synthetic opioids, stimulants, or multiple substances. Finally, performance was evaluated over a single 12-month period; a rolling-origin analysis would provide a stronger assessment of temporal robustness.

Strengths include the comparison of several structurally different forecasting approaches under a common evaluation framework, use of probabilistic performance measures, and presentation of both national and regional trajectories. The forecasts suggest that recent provisional surveillance patterns are compatible with further declines in rolling annual counts, but they do not establish that the decline will persist. Repeated forecast updating, rolling-origin evaluation, and incorporation of revised, substance-specific, and external data are needed before the framework can support stronger operational conclusions.

## 5 CONCLUSION

Using reported provisional NCHS VSRR data, median forecasts from all individual models and ensembles projected lower rolling 12-month-ending overdose death counts by February 2027 nationally and across all four U.S. Census regions. Weighted-ensemble medians projected a 20.5%–21.7% national decline, with larger declines in the Northeast and Midwest than in the South and West. These projections are conditional on the continuation of recent surveillance dynamics. Given the continuing high burden of overdose mortality, the projected declines should not be interpreted as grounds for reducing prevention, evidence-based treatment, naloxone distribution, or harm-reduction efforts.

## Acknowledgments

The authors used generative AI tools to assist with language editing; all content was reviewed and verified by the authors.

## Author contributions

Faharudeen Alhassan: Conceptualization, data curation, formal analysis, methodology, visualization, writing–original draft, writing–review and editing. Hamed Karami: Methodology, formal analysis, validation, writing–review and editing. Robert Bohler: Writing–review and editing. Isaac C. H. Fung: Writing–review and editing. Svenn-Erik Mamelund: Writing– review and editing. Sunmi Lee: Writing–review and editing. Emily N. Peterson: Writing– review and editing. Gerardo Chowell: Conceptualization, supervision, methodology, interpretation, writing–review and editing.

## Data availability

The underlying reported provisional drug overdose death count data used in this study are publicly available from the National Center for Health Statistics Vital Statistics Rapid Release system. The analysis used aggregate, de-identified surveillance data. Processed regional time series and model output files are available at https://github.com/Falhassan123/ Forecasting-of-Drug-Overdose-Deaths/tree/main.

## Ethics statement

This study used publicly available, aggregate, de-identified mortality surveillance data and did not involve human subjects research requiring institutional review board approval.

## 6 Supplementary

**Figure S1:**
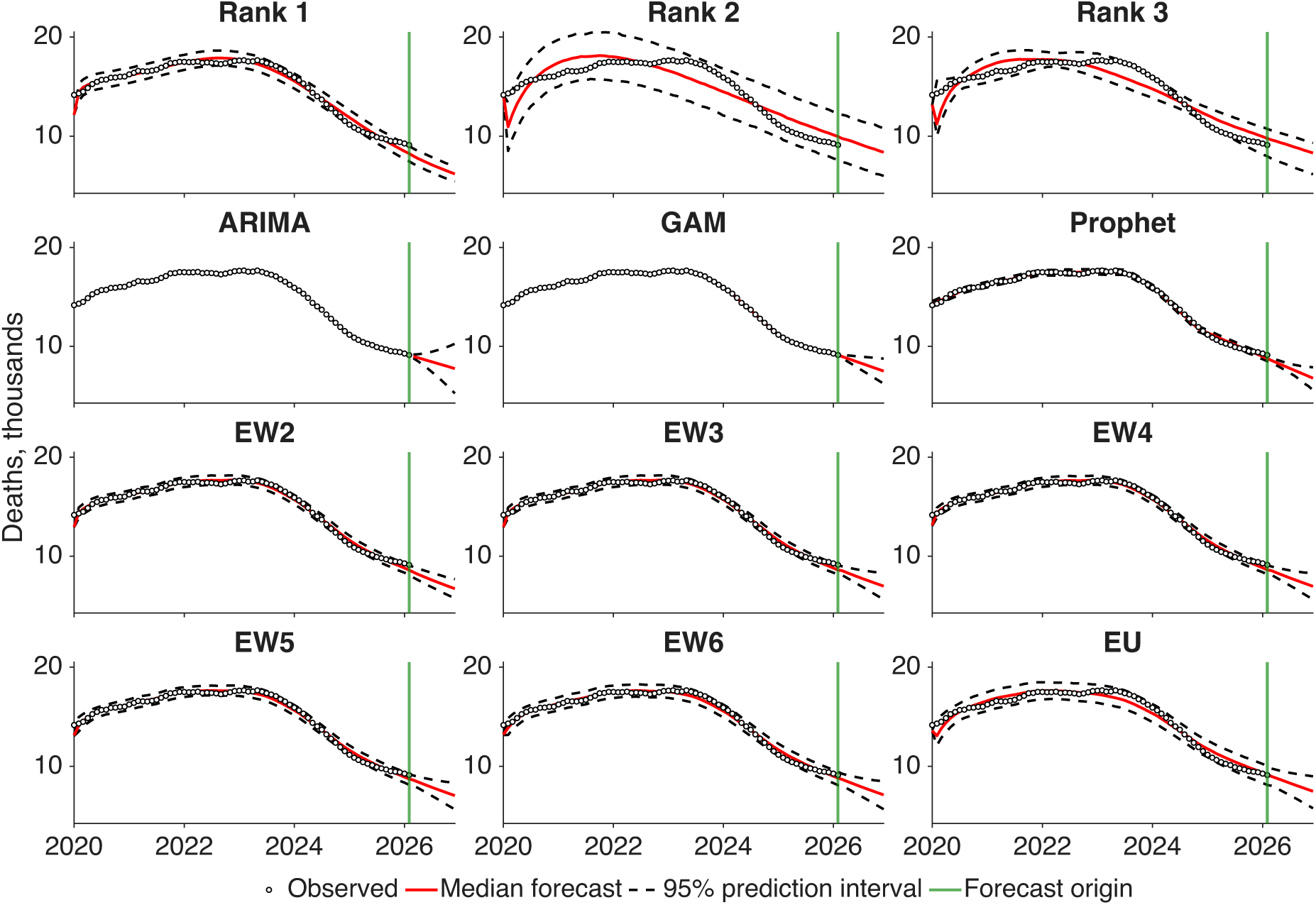
Northeast 12-month-ahead forecasts of rolling annual drug overdose death counts by forecasting model. Observed 12-month-ending overdose death counts from January 2020 through February 2026 are shown as black circles. The vertical green line marks the forecast origin at February 2026, after which models generate forecasts for March 2026 through February 2027. Red curves indicate model-specific median forecasts, and dashed black curves indicate the corresponding 95% prediction intervals. Panels show forecasts from the three AICc-ranked n-sub-epidemic models (Rank 1-Rank 3), ARIMA, GAM, Prophet, WIS-based weighted ensembles EW2–EW6, and the unweighted ensemble EU.

**Figure S2:**
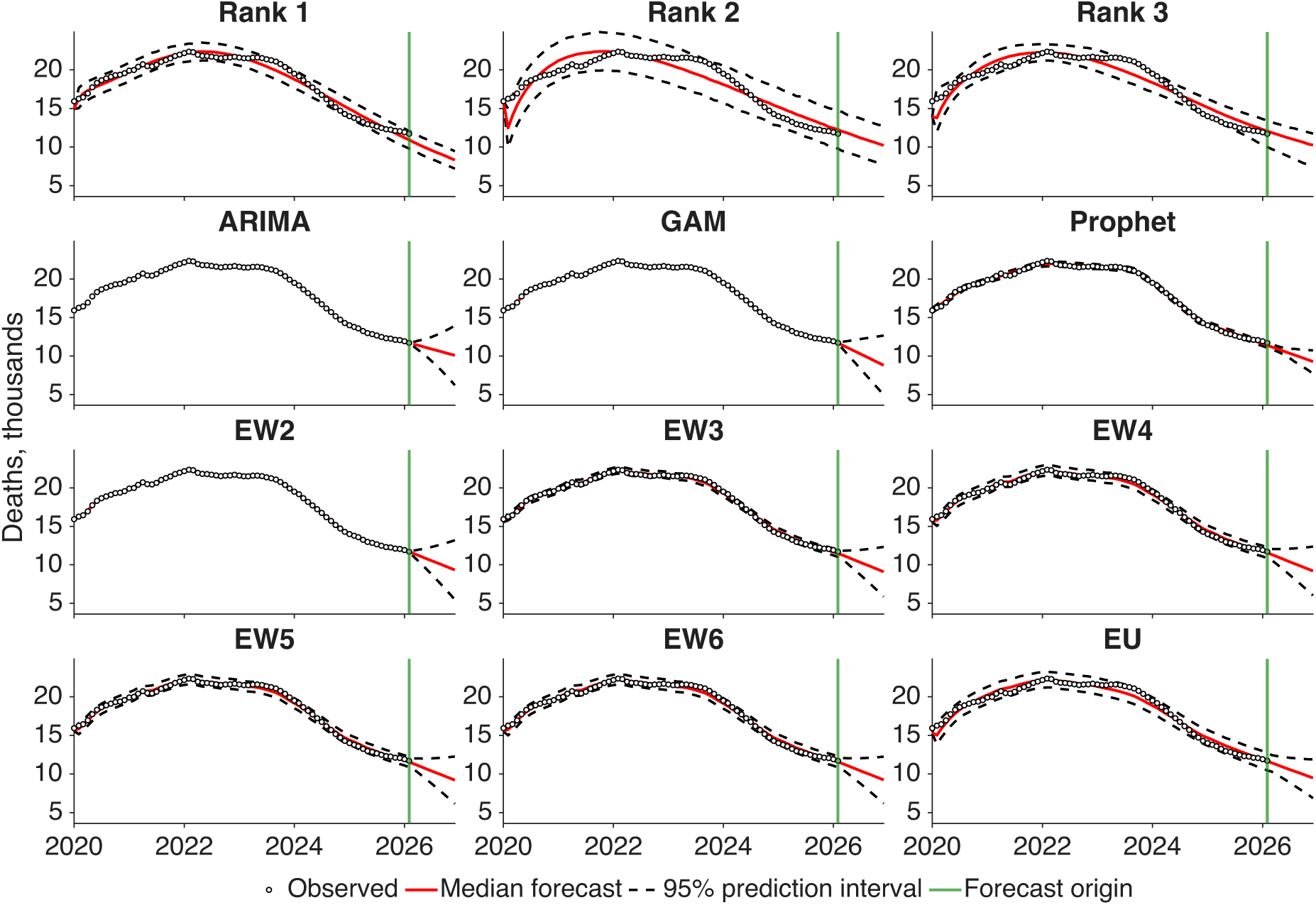
Midwest 12-month-ahead forecasts of rolling annual drug overdose death counts by forecasting model. Observed 12-month-ending overdose death counts from January 2020 through February 2026 are shown as black circles. The vertical green line marks the forecast origin at February 2026, after which models generate forecasts for March 2026 through February 2027. Red curves indicate model-specific median forecasts, and dashed black curves indicate the corresponding 95% prediction intervals. Panels show forecasts from the three AICc-ranked n-sub-epidemic models (Rank 1-Rank 3), ARIMA, GAM, Prophet, WIS-based weighted ensembles EW2–EW6, and the unweighted ensemble EU.

**Figure S3:**
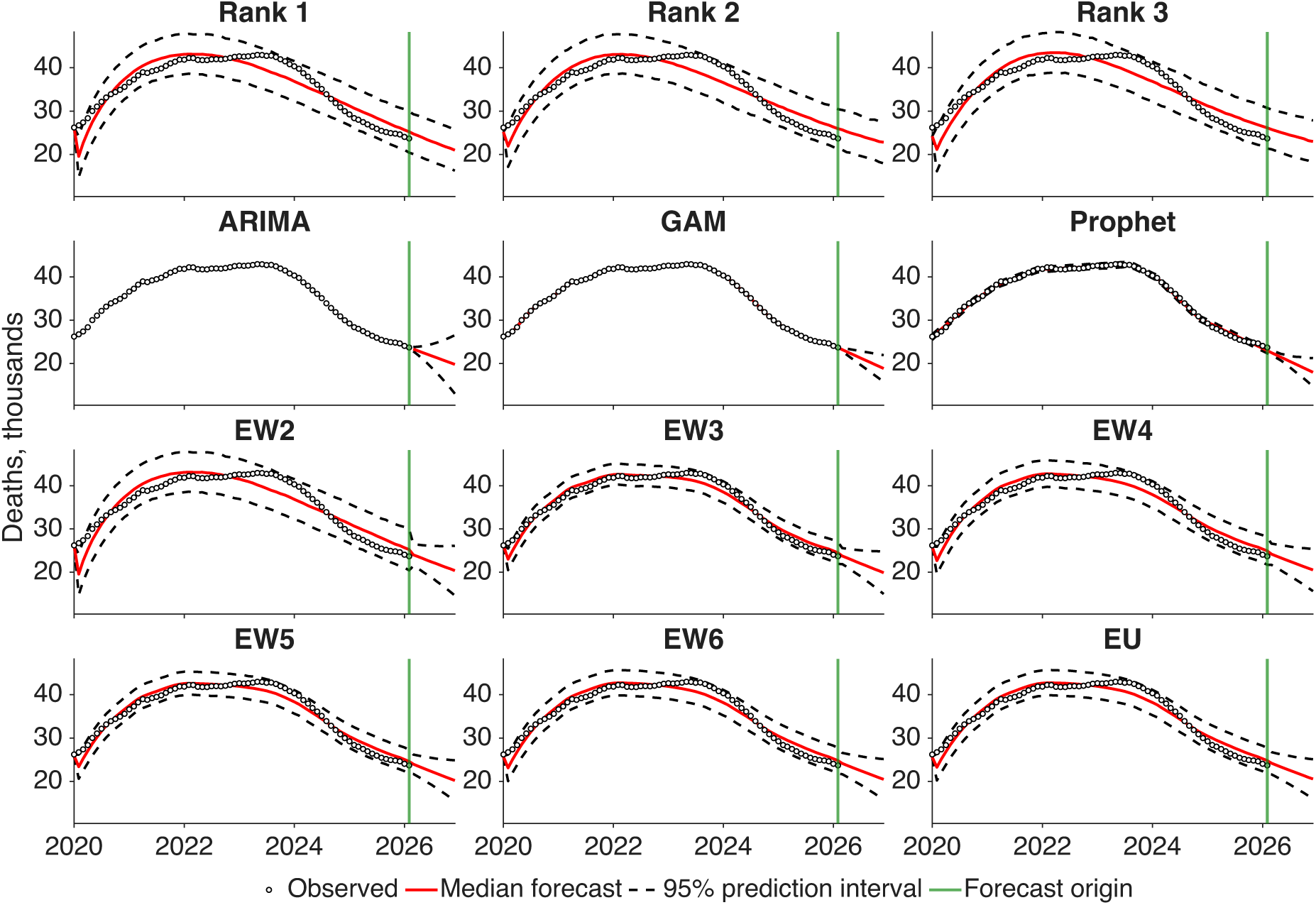
South 12-month-ahead forecasts of rolling annual drug overdose death counts by forecasting model. Observed 12-month-ending overdose death counts from January 2020 through February 2026 are shown as black circles. The vertical green line marks the forecast origin at February 2026, after which models generate forecasts for March 2026 through February 2027. Red curves indicate model-specific median forecasts, and dashed black curves indicate the corresponding 95% prediction intervals. Panels show forecasts from the three AICc-ranked n-sub-epidemic models (Rank 1-Rank 3), ARIMA, GAM, Prophet, WIS-based weighted ensembles EW2–EW6, and the unweighted ensemble EU.

**Figure S4:**
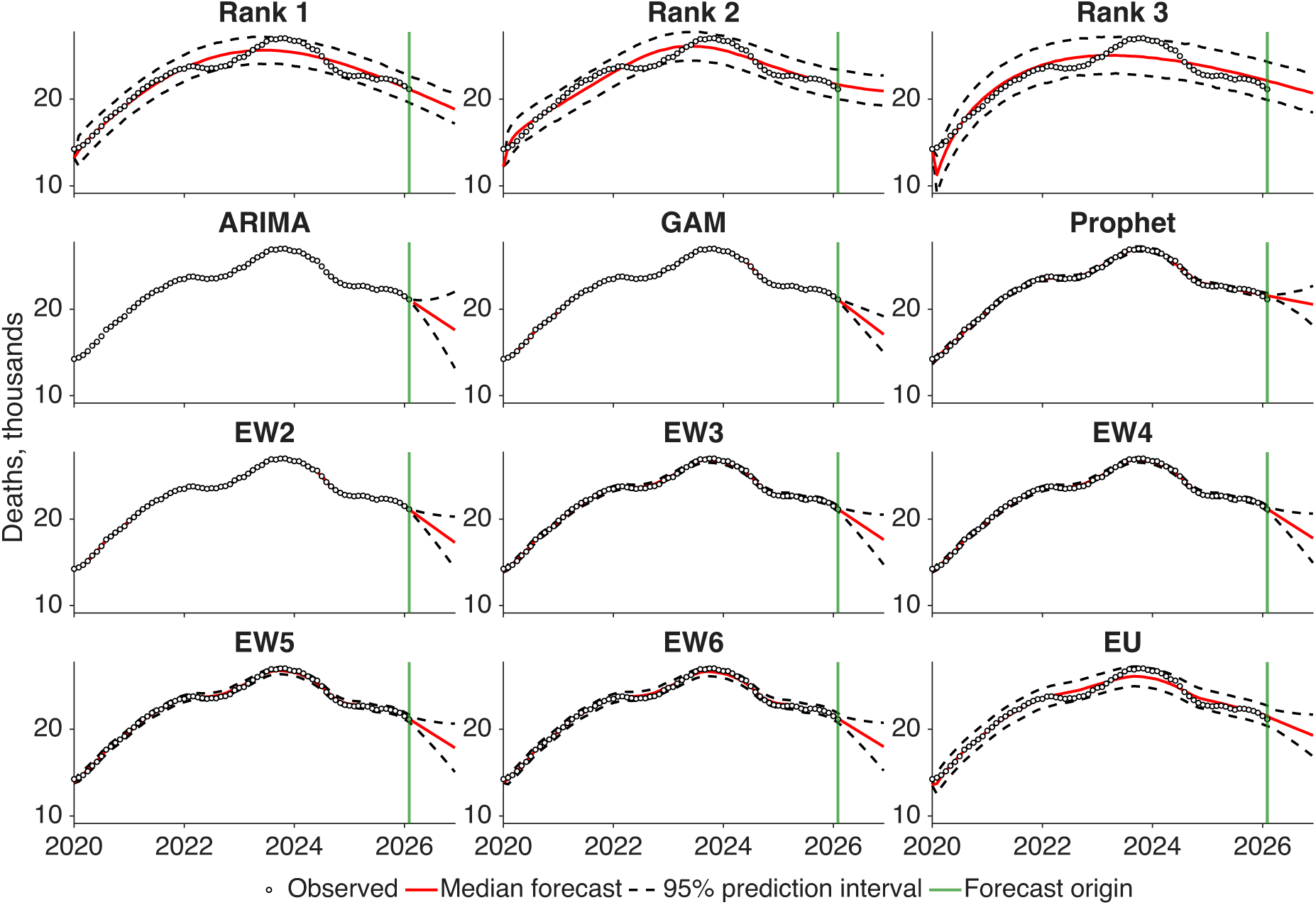
West 12-month-ahead forecasts of rolling annual drug overdose death counts by forecasting model. Observed 12-month-ending overdose death counts from January 2020 through February 2026 are shown as black circles. The vertical green line marks the forecast origin at February 2026, after which models generate forecasts for March 2026 through February 2027. Red curves indicate model-specific median forecasts, and dashed black curves indicate the corresponding 95% prediction intervals. Panels show forecasts from the three AICc-ranked n-sub-epidemic models (Rank 1-Rank 3), ARIMA, GAM, Prophet, WIS-based weighted ensembles EW2–EW6, and the unweighted ensemble EU.

**Table S1:** EW2 inverse-WIS ensemble composition and model weights by geographic series.

| Geographic series | Best ensemble | Weighted WIS summary | Model weights |
| --- | --- | --- | --- |
| National | EW2 | 2.1395 | GAM = 0.5349; ARIMA = 0.4651 |
| Northeast | EW2 | 0.4091 | Rank 1 = 0.6066; GAM = 0.3934 |
| Midwest | EW2 | 0.3397 | GAM = 0.5857; ARIMA = 0.4143 |
| South | EW2 | 1.5299 | ARIMA = 0.5100; Rank 1 = 0.4900 |
| West | EW2 | 0.2567 | GAM = 0.6111; ARIMA = 0.3889 |

EW2 denotes the weighted ensemble constructed from the two individual models with the lowest retrospective WIS within each geographic region. Model weights were calculated using inverse-WIS weighting and normalized to sum to one. The weighted WIS summary is the weighted average of the retrospective WIS values of the two individual models included in the ensemble; it should not be interpreted as the WIS of the pooled ensemble predictive distribution.

